# Longitudinal multi-omic profiling separates persistent from resolving sequelae of breast cancer treatment

**DOI:** 10.64898/2026.09.29.26364331

**Authors:** Heeju Noh, Mary E. Brunkow, Alex V. Carr, Nick Quinn-Bohmann, Lance Pflieger, Samantha M. Megrath, Danielle Burgio, Neil Bailey, Kinsey McCormick, Leroy Hood, Thomas Brown, Charles W. Drescher, Sean M. Gibbons, Nathan D. Price, Erin D. Ellis, Andrew T. Magis, Noa Rappaport

## Abstract

Long-term treatment sequelae shape health and quality of life for years after breast cancer, yet why some women develop them and others recover remains unclear. Here we follow 16 women through chemotherapy and for two years afterwards, with patient-reported symptoms, clinical chemistries, plasma proteomics, metabolomics, and gut microbiome profiles collected across eight visits from a pre-treatment baseline. Sequelae differ in their timing, with some resolving as chemotherapy ends, whereas fatigue, memory problems, and gas bloating can persist, and weight gain and joint pain can appear only afterwards. Weight gain over the second year scales with the cumulative duration of endocrine therapy and is steepest in premenopausal women. The women who remain fatigued at two years are distinguished by a second surge in fatigue scores beginning around 12 months post-chemotherapy rather than by a failure to recover, as well as by elevated plasma INHBB and depleted RAB37. INHBB is also elevated in UK Biobank participants reporting chronic, moderate, or severe fatigue, measured on the same platform. In the gut microbiome, *Faecalibacterium prausnitzii,* a butyrate producer, is depleted in the fatigued group at 24 months post-chemotherapy. Our analysis revealed that post-treatment sequelae follow distinct trajectories, each with its own onset. Markers that track those trajectories point to time windows in which intervention could begin before symptoms become established.

## Introduction

Breast cancer is the most commonly diagnosed cancer among women in the United States, accounting for 32% of all newly diagnosed cancers;^1^ notably, improvements in screening and systemic therapies over the past three decades have shifted the clinical problem from survival toward living well after treatment, with over 4.3 million women living with a history of breast cancer as of January 1, 2025.^2^ For many of them, curative treatment is followed by symptoms that persist for years. Fatigue, cognitive complaints, joint pain, sleep disturbance, and weight gain are among the most frequently reported symptoms, and they carry consequences for quality of life, for the ability to work, and for cardiometabolic risk and cancer recurrence.^3–7^ These symptoms are usually described as a single post-treatment state, yet they differ in when they appear and in how long they last. Some resolve as chemotherapy ends, others persist, and others are first reported months later, at a point when chemotherapy is no longer the only exposure the patient has had. A symptom measured two years after chemotherapy may reflect cytotoxic injury, post-treatment endocrine therapy, a treatment-induced menopausal transition, or a process unrelated to any of these, and the design of most survivorship studies does not allow these variable responses to be separated.

Cancer-related fatigue (CRF) is the best studied of these symptoms and remains among the least well explained. It affects a substantial fraction of patients during treatment and persists in a clinically important minority of survivors years afterwards.^8^ In one longitudinal study, 34% of 763 survivors reported significant fatigue 5 to 10 years after diagnosis and 21% reported it at two assessments several years apart.^9^ CRF is also consistently underreported and undertreated.^10–12^ Large longitudinal cohorts have identified demographic, psychosocial, and treatment-related risk factors, of which pre-treatment fatigue is the most reproducible, which suggests that some of the relevant biology is already present before the first dose.^13^ In the CANcer TOxicity (CANTO) cohort, where predictors measured at diagnosis and treatment exposures were tested together in a validated model of severe fatigue two years later, severe global fatigue was already present in 24.3% of women before treatment began.^14^ Mechanistic work points to inflammatory signalling, with interleukin-6 (IL-6) the most consistent single marker, alongside less consistently demonstrated contributions from hypothalamic-pituitary-adrenal axis function, mitochondrial metabolism, and sleep.^15,16^ Longitudinal studies show more consistent inflammatory associations than cross-sectional ones.^17^ Most molecular studies of fatigue in this population have relied on targeted cytokine panels, candidate genetic variants, or metabolomics in small cross-sectional samples. It therefore remains unclear when the women who develop persistent fatigue begin to differ from those who recover: at diagnosis, at the end of treatment, or later still.

Weight gain after treatment is similarly common, and its cause is similarly unsettled. Body weight increases during chemotherapy by 2.7 kg, on average, across 25 studies and 2,620 women, with heterogeneity high enough (I^2^ = 94.2%) that the average conceals wide variation between patients.^18^ Survivors are more likely to gain weight than age-matched women without a history of cancer, and the gain carries measurable risk, since higher body weight in this population is associated with lymphedema, cardiometabolic disease, and cancer recurrence.^7^ Endocrine therapy is frequently implicated in this weight gain, but randomized comparisons do not support a direct pharmacological effect: across the Arimidex, Tamoxifen, Alone or in Combination (ATAC) trial and the International Breast Cancer Intervention Studies I and II (IBIS-I and IBIS-II), mean 12-month weight change did not differ between tamoxifen and placebo, between anastrozole and placebo, or between anastrozole and tamoxifen.^19^ Observational cohorts instead find weight gain concentrated in a subset of patients rather than distributed across all of them, with younger and premenopausal women over-represented among those who gain weight.^20^ Body weight is also a coarse endpoint for the underlying physiology. Women receiving aromatase inhibitors gain fat mass and lose lean mass over 18 months, changes that a single weight measurement cannot capture,^21^ and we have previously shown that multi-omic signatures of body mass index (BMI) track metabolic heterogeneity and response to lifestyle intervention more closely than measured BMI.^22^

The gut microbiome is a plausible contributor to both phenotypes and has rarely been measured in breast cancer cohorts. Chemotherapy and antibiotics alter microbial composition and metabolic output, and microbially derived short-chain fatty acids (SCFAs) influence host energy balance and inflammation.^23,24^ Depletion of butyrate-producing capacity has been reported in myalgic encephalomyelitis and chronic fatigue syndrome,^25^ and reduced dominance of butyrate-producing organisms has been associated with poorer health and survival outcomes in older cohorts.^26^ The sparse sampling of cohorts large enough to model risk factors, the cross-sectional design of most molecular studies, and the absence of pre-treatment baselines limit our mechanistic understanding of variable responses to treatment. To address this, the Institute for Systems Biology and the Providence Swedish Cancer Institute conducted the Surviving Through Breast Cancer (VIBE) study, a prospective longitudinal cohort of women receiving chemotherapy for breast cancer, profiled from before their first dose through two years after treatment ended. Herein, we analyze patient-reported symptoms, clinical chemistries, plasma proteomics, untargeted metabolomics, and gut microbiome composition across eight timepoints, and use the density of sampling to separate the sequelae that resolve with treatment from those that persist or emerge post-treatment, and to identify the clinical, dietary, and molecular factors that accompany them.

## Results

### Study design and cohort description

We studied 16 women with breast cancer (mean age 45.1 ± 11.5 years) enrolled in the VIBE study, a prospective longitudinal cohort followed from before chemotherapy to 24 months after its completion **(Fig. 1a)**. The cohort divides evenly by menopausal status, with 8 premenopausal and 8 postmenopausal participants, and age is distributed around the menopausal transition rather than evenly across the range **(Fig. 1b)**. Ten patients had HR+/HER2-disease, four had HR+/HER2+ disease, and two had triple-negative breast cancer (TNBC) **(Fig. 1c)**. All patients underwent (neo)adjuvant chemotherapy and surgery, most received adjuvant radiation therapy, and 12 of the 16 patients (75%) proceeded to endocrine therapy, which for most of them was still ongoing at the end of follow-up **(Fig. 1d)**. Patient-reported outcomes, blood chemistries, proteomics, metabolomics, and gut microbiome profiling were collected at baseline, during chemotherapy, at the end of chemotherapy, and at 3, 6, 12, 18, and 24 months post-chemotherapy, with questionnaires and clinical chemistries collected at every visit and the omics assays concentrated at baseline, the end of chemotherapy, and 12 and 24 months **(Fig. 1e)**. We began by characterizing which side effects persist beyond treatment, then examined the clinical, dietary, and molecular factors that accompany the long-term fatigue and weight gain phenotypes in this cohort.

**Figure 1.**
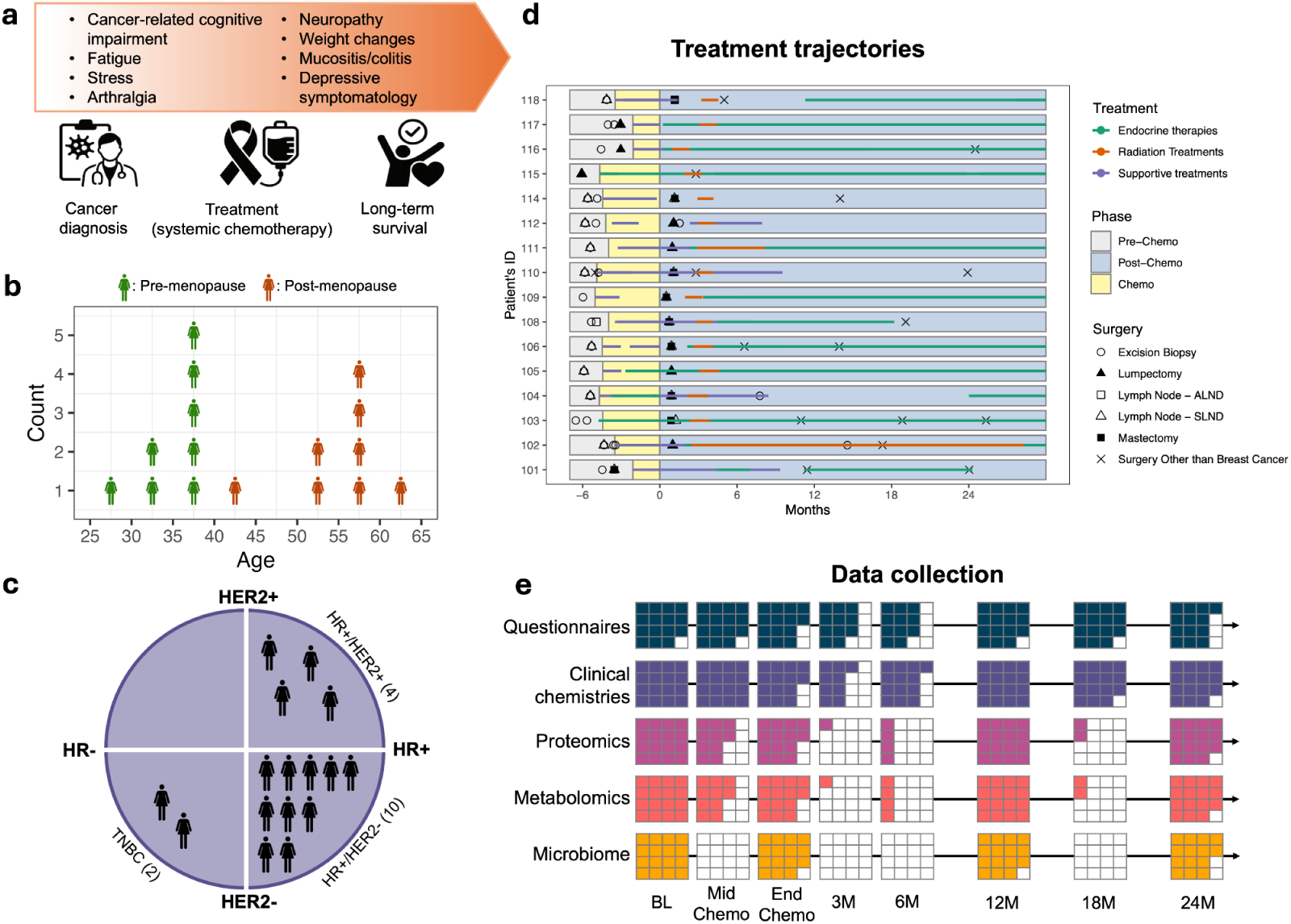
Study design, cohort characteristics, and longitudinal sampling timeline. **(a)** Study schematic. The VIBE study follows women with breast cancer from diagnosis through systemic chemotherapy into long-term survivorship, tracking the treatment-related adverse effects listed, from a pre-chemotherapy baseline through 24 months post-chemotherapy. **(b)** Age distribution of the 16 participants, binned in 5-year intervals, colored by menopausal status: premenopausal (n=8, green) and postmenopausal (n=8, orange). **(c)** Tumor subtypes across the cohort (n=16). HR: hormone receptor; HER2: human epidermal growth factor receptor 2; TNBC: triple-negative breast cancer. **(d)** Swimmer plot illustrating individual patient treatment profiles (n=16). The x-axis indicates timeline relative to the completion of chemotherapy (t=0). Shaded background regions denote pre-treatment (grey), active chemotherapy (yellow), and post-chemotherapy (blue) phases. Line colors and lengths indicate therapy type and duration in months (green: endocrine therapy; orange: radiation therapy; purple: supportive therapies). Point symbols specify surgical interventions: open circle, excision biopsy; filled triangle, lumpectomy; open square, axillary lymph node dissection (ALND); open triangle, sentinel lymph node dissection (SLND); filled square, mastectomy; cross mark, non-breast surgery. **(e)** Matrix of multi-omics and clinical sample collection across time points (BL: baseline; MidChemo: mid-chemotherapy; EndChemo: completion of chemotherapy; 3M-24M: 3 to 24 months post-chemotherapy) for self-reported questionnaires (dark blue), clinical chemistries (purple), proteomics (pink), metabolomics (orange), and gut microbiome (yellow); white indicates missing data.

### Ethics statement

The current study was conducted with de-identified data of the participants who had consented to the use of their anonymized data in research. Procedures were run under the WCG Institutional Review Board, formerly Western IRB (IRB Tracking #20171513; Study #1176924). This research also includes analysis of the UK Biobank Resource under Application Number 48049.

### Fatigue and memory problems persist after chemotherapy ends

To identify the side effects that persist beyond treatment, we analyzed basic measurements such as weight, body temperature, and blood pressure alongside self-reported adverse events. We first converted ordinally lcategorized variables representing the frequency or severity of side effects into numerical values corresponding to grades on a normalized scale from 0 to 5, then averaged variables belonging to the same side-effect group (see Methods), and compared the resulting scores during and after chemotherapy to each patient’s baseline **(Fig. 2)**. Weight increased after chemotherapy (FDR-adjusted p_post_=0.0069 by two-tailed, one-sample Wilcoxon signed-rank test), while body temperature (ppost=0.00096) and heart rate (ppost=0.0089) fell over the same period; BMI, blood pressure (BP), and respiration rate either did not change significantly or exhibited a median score change of zero across both periods (**Fig. 2; Supplementary Fig. 1**). Among the 41 self-reported side-effect groups, gas bloating (pchemo=0.0024; ppost=0.0002), fatigue (p_chemo_=0.0011; p_post_=0.0019), and memory problems (p_chemo_=0.0011; p_post_=0.0010) were elevated both during and after chemotherapy, whereas joint pain reached significance only after chemotherapy (p_post_=0.0003). Hot flushes (p_chemo_=0.0086), eye problems (p_chemo_=0.0024), difficulty concentrating (p_chemo_=0.0024), and pain-related side effects (p_chemo_=0.0024) were elevated during chemotherapy and returned toward baseline afterwards, while breast tenderness was lower than baseline in both periods (p_chemo_=0.0011; p_post_=0.0006), and lower stress (p_post_=0.0003) and higher activity (p_post_=0.0002) were achieved after chemotherapy. These results indicate that the symptom burden of treatment separates into at least two temporal patterns, one that resolves as chemotherapy ends and one that persists or emerges afterwards.

**Figure 2.**
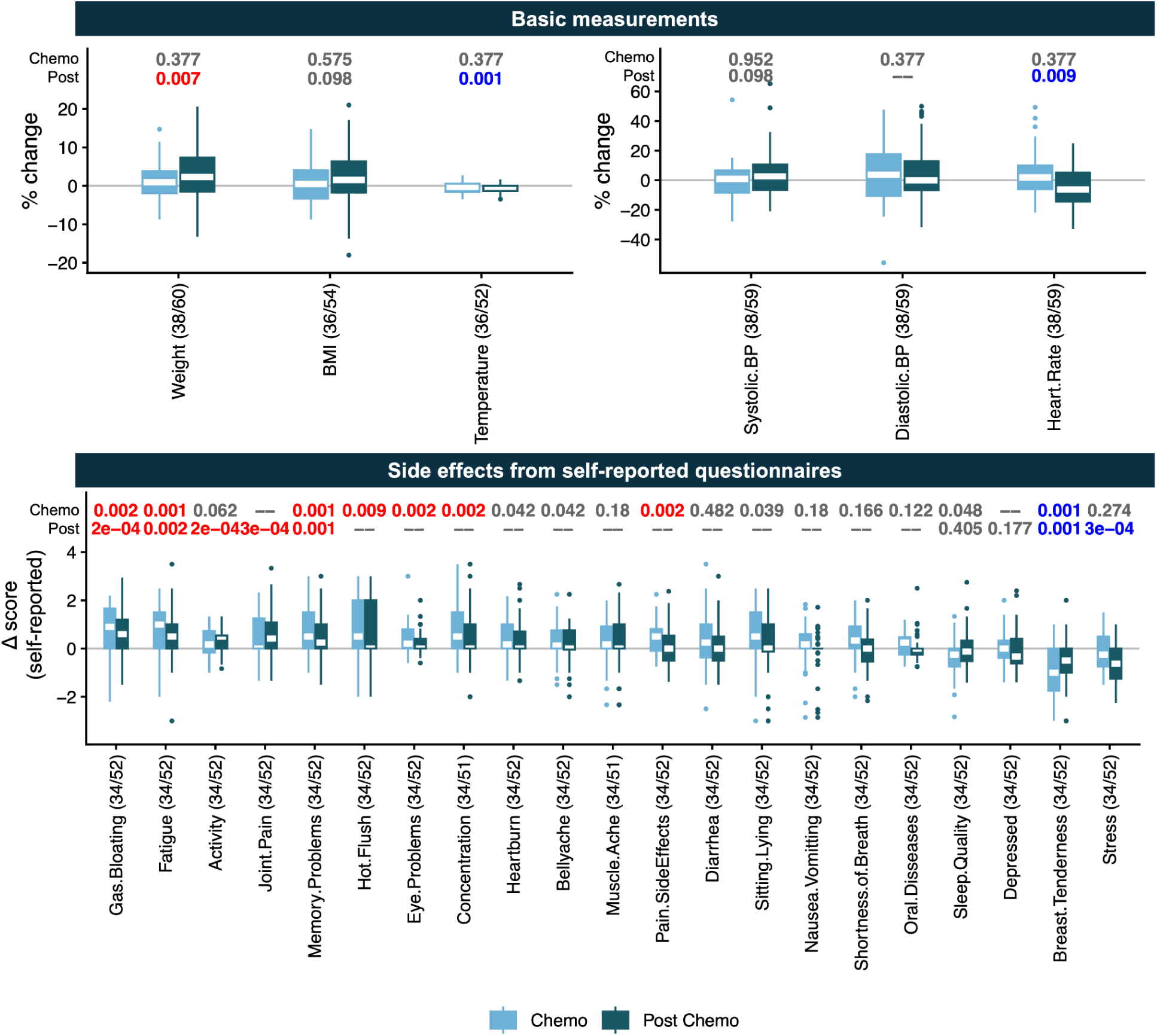
Dynamic changes in body measurements and side-effect scores during and post-chemotherapy. Panels display percent changes in body measurements (top) and score changes across 21 self-reported side-effect groups (bottom) relative to baseline during chemotherapy (light blue; treatment to 3 months post-last dose) and post-chemotherapy (dark blue; 3–24 months post-treatment). Side-effect scores were normalized (0–5 scale; Methods). Box plots show the median (center line), upper and lower quartiles (box limits), and whiskers extending to 1.5×IQR(interquartile range) from the hinges. Individual points beyond the whiskers denote outliers. Statistical significance was assessed using two-tailed, one-sample Wilcoxon signed-rank tests. Sample sizes for each test are annotated on the x- axis labels as "side effect (n[Chemo]/n[PostChemo])". Top annotations show FDR-adjusted p-values (q<0.01) for Chemo (during chemotherapy) and Post (post-chemotherapy), where red denotes significant increases and blue denotes significant decreases. Variables with a median of 0 were excluded from statistical testing ("--") and omitted if zero in both phases (full set of 41 variables shown in Supplementary Figure 1).

### Weight gain tracks endocrine treatment duration

Weight gain and fatigue were the two side effects that persisted or emerged in our cohort once treatment ended. Both carry long-term health consequences for survivors^27,28^, and therefore we investigated parameters that may have affected these two side effects after chemotherapy. To do so, we fit generalized linear models (GLM)^29^ to patients’ treatment and tumor information, and generalized estimating equations (GEE)^30^ to 28 self-reported diet-related variables over time, using percent weight change and fatigue score change as response variables. For weight gain, longer duration of endocrine therapy (p=0.0005, FDR-adjusted) was significantly associated with weight gain after chemotherapy, while higher cheese intake (p=0.016, FDR adjusted) was inversely associated with weight gain after chemotherapy, out of 13 treatment and tumor variables and 28 dietary variables tested (**Fig. 3a**). Tumor type (HR+ or TNBC) was also nominally associated (p= 0.064, FDR-adjusted) with weight gain, as TNBC patients did not receive endocrine therapy, while age, baseline weight, and BMI showed no association in this model. Notably, we measured total body weight, which does not resolve the underlying shifts in fat and lean mass or in metabolic state. Patients with limited endocrine exposure (total duration <10 months), a threshold selected based on an observed bimodal distribution with a gap near 10 months, remained weight-stable through 12 months post-chemotherapy and then lost weight, reaching a median of −5% relative to baseline weight at 24 months post-chemotherapy, whereas premenopausal and postmenopausal groups with ≥10 months of total endocrine treatment exposure gained weight steadily and separated from each other after 12 months post-chemotherapy, with premenopausal patients reaching a median weight gain of nearly 9% and postmenopausal patients of nearly 5% at 24 months post-chemotherapy **(Fig. 3b)**. Among measurements collected between 12 and 24 months post-chemotherapy, duration of endocrine therapy was positively correlated with weight gain in premenopausal patients (r^2^ = 0.518, p = 9.5×10^-5^), and more weakly so in postmenopausal patients (r^2^ = 0.138, p = 0.042), and the weight gain in premenopausal patients was significantly greater than in postmenopausal patients among those who received ≥10 months of therapy (p = 0.0087 by two-tailed Mann-Whitney U-test) **(Fig. 3c)**. Because menopausal status is associated with age and body weight in this cohort, we repeated the comparison stratifying by age (≥45 vs. <45 years) and by baseline body weight (≥80 kg vs. <80 kg), and observed the same pattern under both stratifications (p = 0.00018 and p = 1.6×10^-5^, respectively) **(Supplementary Fig. 2a and 2b)**. Under both stratifications, the association with endocrine duration was stronger in the younger group (r^2^ = 0.528) and the lower-weight group (r^2^ = 0.467) than in their counterparts. In the older group it did not reach significance (r^2^ = 0.093, p = 0.097), while in the heavier group it remained significant (r^2^ = 0.380, p = 0.0013). None of these factors were associated with weight gain when short-and long-term endocrine treatment groups were analyzed together **(Fig. 3a)**, indicating that age, weight, and menopausal status largely separate the same patients rather than acting as independent predictors.

**Figure 3.**
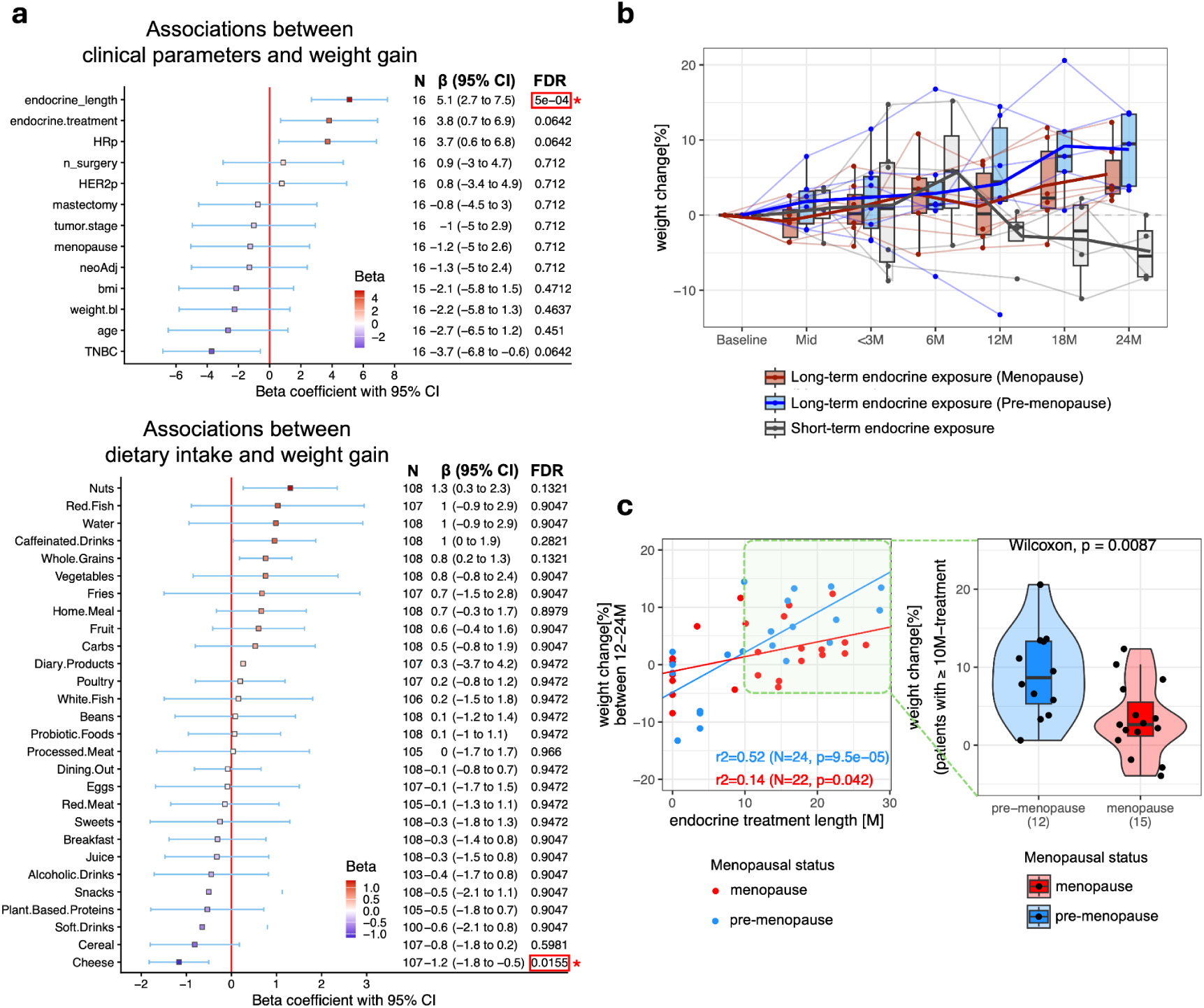
Clinical, dietary, and treatment factors associated with post-chemotherapy weight gain. **(a)** Forest plots displaying estimated β coefficients from a GLM evaluating tumor and treatment variables (top) and a GEE evaluating time-series dietary variables collected after chemotherapy (bottom), with percentage weight change at 24 months as the response. Positive and negative β coefficients are represented by red (+) to blue (-) color scales in squares, with white representing zero. Error bars denote 95% confidence intervals (CIs). Sample sizes (N), β coefficients (95% CIs), and FDR-adjusted p-values are annotated on the right side of each panel. Significant p-values are highlighted with a red box. **(b)** Trajectories of percent weight change from baseline to 24 months post-chemotherapy for three groups: postmenopausal patients with ≥10 months of endocrine therapy (red, n=6), premenopausal patients with ≥10 months (blue, n=6), and patients with <10 months (grey, n=4). Box plots show the distribution within each group at each timepoint, bold lines connect the group medians, and faint lines show individual participants. The dashed line marks no change from baseline. **(c)** Left, scatter plot of endocrine treatment length (M, months) against percent weight change, pooling the 12M, 18M, and 24M post-chemotherapy timepoints, colored by menopausal status (premenopausal, blue; postmenopausal, red), with linear fits and r^2^, sample size (N), and p values shown within each group. Shading marks the patients and interval selected for the comparison at right. Right, violin plot of percent weight change between 12 and 24 months post-chemotherapy in postmenopausal (red) and premenopausal (blue) patients who received ≥10 months of endocrine therapy, compared by two-tailed Mann-Whitney U test. Violins show the density with an inset box plot and individual points overlaid. Box plots show the median (center line), upper and lower quartiles (box limits), and whiskers extending to 1.5×IQR(interquartile range) from the hinges.

### INHBB and RAB37 distinguish patients with persistent fatigue

We repeated the above analysis to identify factors associated with persistent fatigue after chemotherapy. At 24 months post-chemotherapy, four patients exhibited a markedly higher increase in fatigue score relative to the remaining patients. Comparing these patients (fatigue group, n=4) with the rest of the participants (non-fatigue group, n=12) using GLM and GEE models revealed no factors that maintained statistical significance after multiple hypothesis correction; however, probiotic food intake produced the largest dietary coefficient and showed a nominal positive association with fatigue (p=0.0886, FDR-adjusted) (**Fig. 4a**). Patients lacking fatigue score data at 24 months were excluded. Consequently, we shifted our focus toward identifying molecular signatures of long-term fatigue. Fatigue scores rose in both groups during chemotherapy and fell back toward baseline within three months of its end, after which the two groups diverged, with the fatigue group rising again from 12 months onward **(Fig. 4b)**. The long-term phenotype therefore reflects a late second increase rather than a failure to recover from treatment. Among the multi-omic features tested, the proteins INHBB and RAB37 were positively and negatively associated with long-term fatigue, respectively **(Fig. 4c)**. Trajectories of normalized abundances over time demonstrated that INHBB levels were significantly elevated (p=0.011 by a two-tailed, two-sample Mann-Whitney U test) in the fatigue group at postChemo (12-24 months post-chemotherapy) **(Fig. 4d)**, whereas RAB37 was significantly depleted both at endChemo (from the end of chemotherapy up to 3 months post-chemotherapy) (p=0.012) and at postChemo (p=0.00044) **(Fig. 4e)**. The two markers separated the groups at different times, with RAB37 lower in the fatigue group from mid-chemotherapy onward and INHBB separating the groups only after treatment ended. To evaluate whether these markers are associated with fatigue outside the treatment setting, we compared plasma INHBB and RAB37 levels across fatigue subgroups in the UK Biobank,^31^ measured on the same Olink platform used in VIBE. INHBB was elevated in participants reporting chronic fatigue (i.e. more than 6-month persistent or recurrent tiredness; p = 2.83×10^-7^ by by a two-tailed, two-sample t-test) and in those reporting moderate (p = 2.90×10^-7^) or severe (p = 0.00192) fatigue over the past week, but not in those reporting mild fatigue (p = 0.12), and was also elevated in participants reporting more than 12 weeks of mild fatigue relative to those reporting less than 2 weeks (p = 0.0090), indicating that the association tracks fatigue duration as well as severity. RAB37 did not differ significantly in any UK Biobank comparison **(Supplementary Table 1)**. UK Biobank fatigue is not treatment-related fatigue after breast cancer, so these comparisons provide cross-context support for an INHBB association rather than replication of the VIBE finding.

**Figure 4.**
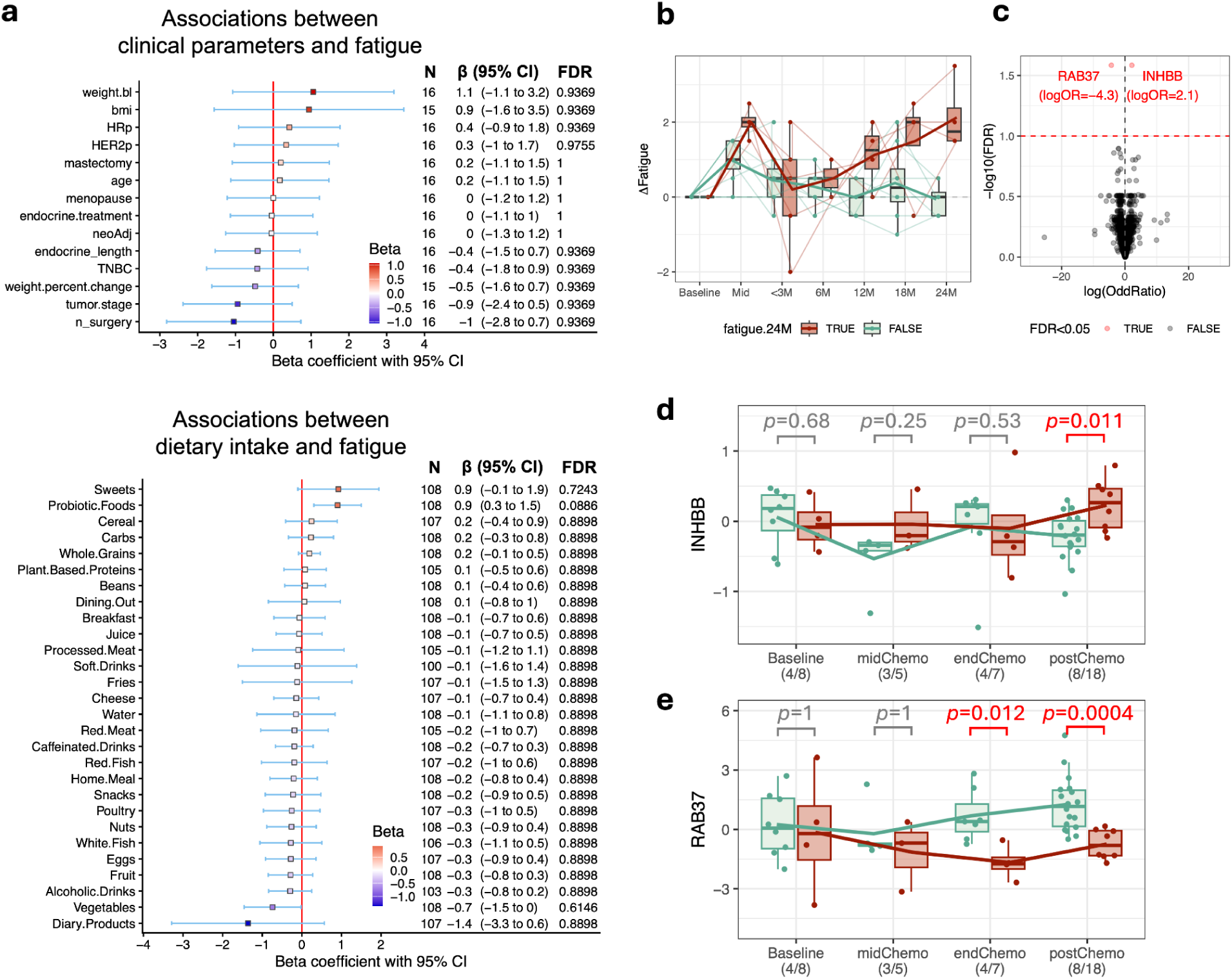
Clinical, dietary, and molecular correlates of post-chemotherapy fatigue. **(a)** Forest plots displaying estimated β coefficients generated in the same manner as Figure 3A. A binary response distinguishing fatigue and non-fatigue groups was used as the outcome. The fatigue group was defined as four patients who showed an outlying increase in fatigue at 24 months post-chemotherapy (Methods). **(b)** Longitudinal trajectories of baseline-normalized fatigue scores comparing fatigue and non-fatigue groups. The fatigue group comprises the 4 patients whose fatigue score increased by more than 1.5 at 24 months post-chemotherapy; the non-fatigue group comprises the 8 patients with a change of 1.5 or less. Box plots show the distribution within each group at each timepoint, bold lines connect the group medians, and faint lines show individual participants. **(c)** Volcano plot of log(odds ratio) vs.-log_10_(FDR) from GEE analysis of multi-omics fatigue signatures (blood chemistry, metabolomics, and proteomics). Statistically significant analytes (FDR<0.05) are highlighted in red. **(d, e)** Trajectories of normalized INHBB (d) and RAB37 (e) abundance at baseline, mid-chemotherapy (midChemo), end of chemotherapy (endChemo: 0-3M), and post-chemotherapy (postChemo: 12-24M) time points. Box plots show the median (center line), upper and lower quartiles (box limits), and whiskers extending to 1.5×IQR(interquartile range) from the hinges. Individual data points are overlaid. P-values from two-tailed Mann-Whitney U tests comparing the fatigue and non-fatigue groups at each timepoint are annotated above the corresponding box plots, with statistically significant differences (p<0.05) highlighted in red. Sample sizes for each test are annotated on the x-axis labels as "timepoint (n[fatigue]/n[non-fatigue])”.

### Depletion of *F. prausnitzii* in patients with persistent fatigue

Butyrate is a gut microbiome-derived short-chain fatty acid (SCFA) known to influence frailty and weight dynamics.^32–34^ To assess whether the fatigue and weight gain phenotypes were accompanied by a shift in gut microbial metabolic capacity, we employed Microbial Community-Scale Metabolic Modeling (MCMM)^35^ to predict butyrate production in the VIBE gut microbiome samples under a simulated personalized diet constraint (see Methods). Permutation-based analysis of predicted butyrate production revealed a progressive decrease in flux differences between groups (Δflux=butyrate_fatigue_−butyrate_non-fatigue_) over time. Specifically, butyrate production in the fatigue group was significantly lower than in the non-fatigue group at 24 months post-chemotherapy (mean Δflux=−0.95 and p=0.044, FDR-adjusted by two-tailed, one-sample Wilcoxon signed-rank test) **(Supplementary Fig. 3)**. We therefore examined three of the most prevalent butyrate producers of the human gut, *Faecalibacterium prausnitzii*, *Roseburia intestinalis*, and *Eubacterium rectale*.^36,37^ At 24 months post-chemotherapy, lower relative abundance of *F. prausnitzii* was significantly associated with fatigue in logistic regression analysis (p = 0.0316), while *R. intestinalis* and *E. rectale* did not differ between the groups. *F. prausnitzii* was also lower in the fatigue group at baseline, though not significantly (p = 0.142), so we cannot rule out a difference that predates treatment. This association does not survive correction across all detected taxa and holds only under a targeted analysis restricted to these three butyrate producers, so we report it as a candidate finding. Its direction is consistent with reported deficiencies of *F. prausnitzii* and of butyrate-producing capacity in individuals with chronic fatigue syndrome.^25^

## Discussion

Improvements in detection and therapy have produced a growing population of breast cancer survivors, estimated at 4.3 million in the United States.^2^ Fatigue and weight gain persist for years, eroding quality of life and adding to the cost of survivorship care,^3,38^ yet the processes separating patients who develop these sequelae from those who do not remain poorly characterized. Here we followed 16 women through chemotherapy and two years afterwards, integrating patient-reported outcomes with clinical chemistries, proteomics, metabolomics, and gut microbiome profiles. The major findings were: 1) side effects were heterogeneous in their timing, with hot flushes and difficulty concentrating resolving as chemotherapy ended, fatigue and memory problems persisting, and weight gain and joint pain appearing only afterwards; 2) weight gain over the two years following chemotherapy tracked cumulative endocrine therapy duration, particularly in premenopausal women; 3) persistent fatigue appeared as a post-chemotherapy sequelae, with elevated plasma INHBB (also associated with chronic fatigue in the UK Biobank) and depleted RAB37; and 4) the abundance of *F. prausnitzii*, a predominant butyrate-producing gut bacterium, was less prevalent in the fatigue group at 24 months post-chemotherapy.

The heterogeneous onset of side effects indicates that factors beyond chemotherapy are involved. Hot flushes, eye problems, difficulty concentrating, and pain-related complaints rose during chemotherapy and returned toward baseline once therapy ended, whereas fatigue, memory problems, and gas/bloating remained elevated post-treatment, and weight gain and joint pain reached significance only after treatment. Stress fell and physical activity rose, consistent with recovery from the acute burden of treatment. The likely contributor to the late-emerging symptoms is endocrine therapy, which 12 of the 16 patients received, with most continuing hormone therapy up through the end of follow-up. Joint pain is a well-established adverse effect of aromatase inhibitors, reported in about half of treated women,^39^ suggesting that symptoms at 24 months cannot be attributed to chemotherapy alone. Separating transient from persistent fatigue likewise requires a pre-treatment measurement, and pre-treatment fatigue is among the strongest reported predictors of persistent post-treatment fatigue.^14,40,41^

Weight gain over the second year was positively associated with cumulative endocrine therapy duration. Randomized trials show comparable 12-month weight change across ATAC, IBIS-I, and IBIS-II, whether tamoxifen was compared with placebo, anastrozole with placebo, or anastrozole with tamoxifen.^19^ Larger studies find post-diagnosis weight gain concentrated in a subset, with premenopausal status among the factors identified.^20^ In our cohort, neither menopausal status nor its correlates, age and baseline weight, independently explained the year 2 weight changes when short-and long-term exposure groups were analyzed together. Stratifying by menopausal status and cumulative endocrine duration suggests that susceptibility depends on the interaction between the two rather than on treatment status alone, pointing to the treatment-induced hormonal transition as a driver, consistent with prior evidence.^28,42^ The underlying physiology differs by menopausal status. In 43 premenopausal women followed with DXA from before chemotherapy to 12 months, the 30 who developed chemotherapy-induced ovarian failure gained a median 1.8 kg of truncal fat and lost 0.6 kg of truncal lean mass, changes not seen in those who retained ovarian function.^43^ Among postmenopausal women, aromatase inhibitors increase fat mass and reduce lean mass over 18 months, shifts that total body weight cannot capture.^21^ The trajectories reported here therefore describe body mass, and the underlying changes in body composition may be larger. Cheese intake was the only dietary variable significantly and inversely associated with weight gain, consistent with previous reports demonstrating an inverse association between cheese consumption and weight gain^44^.

^(Pedersini^Fatigue rose in all participants during chemotherapy and returned toward baseline within three months, after which a subset showed a secondary rise around 12 months that persisted to 24 months. This trajectory suggests that persistent fatigue arises from shifts occurring after recovery rather than from a continuation of acute treatment effects. That division has support elsewhere: in a prospective cohort, the development of fatigue followed therapy whereas its persistence was driven by conditions present beforehand, including depression, pain, and lifestyle factors.^45^ In REQUITE,^46^ 1,443 women followed for two years showed fatigue rising throughout treatment and then returning to baseline following treatment,^41^ consistent with the aggregate pattern in our non-fatigue group. Averaging across a cohort of that size can conceal a subgroup whose fatigue rises again after recovery. IL-6 is widely regarded as the most consistent single biomarker of CRF,^15,16^ yet INHBB and RAB37 were the features separating the fatigue group in our data. Plasma INHBB rose only in the 12 to 24 month window, whereas RAB37 fell both at the end of chemotherapy and across that same window. In the UK Biobank, which uses the same Olink platform, plasma INHBB was likewise elevated among participants reporting chronic, moderate, or severe fatigue, and tracked the duration of mild fatigue. INHBB forms a subunit of activin B, itself a reported biomarker of chronic fatigue,^47^ providing a potential mechanistic link. RAB37 is a Rab family GTPase involved in membrane trafficking and vesicle secretion^48^, suggesting a potentially new mechanistic link, although it did not differ significantly in UK Biobank.

*F. prausnitzii*, a prevalent butyrate-producing bacterium,^36^ was depleted in the fatigue group at 24 months. Because the association did not survive FDR correction across all taxa, we present it as a candidate result that will have to be corroborated with a larger sample size. At the same timepoint, the median predicted butyrate production was also significantly lower in the fatigue group. Reduced butyrate-producing capacity has been reported in myalgic encephalomyelitis and chronic fatigue syndrome,^25^ making *F. prausnitzii* a reasonable prior for a larger study.

There were several limitations to our study. The cohort of 16 women constrains statistical power and generalizability, and 4 lacked fatigue scores at 24 months, restricting that stratification to 12 (fatigue, n=4; non-fatigue, n=8), so few features retained significance after FDR correction. Fatigue groups were defined empirically by identifying participants with outlier fatigue score increases at 24 months post-chemotherapy compared to the rest of the cohort, rather than a validated cutoff, and the UK Biobank comparison is not specific to treated cancer patients, so confirmation requires an independent treated cohort. Body composition was not measured, and endocrine regimens differed between the menopausal groups: all postmenopausal participants received an aromatase inhibitor, while of the six HR+ premenopausal women, four received tamoxifen and two letrozole with ovarian function suppression. Weight effects do not appear to differ between aromatase inhibitors and tamoxifen,^19^ but we cannot rule out agent-specific effects alongside the physiological transition. The cohort was single-site and demographically homogeneous, and the regression models assume linear relationships between dietary variables and outcomes. The side effects and biomarkers identified here should therefore be treated as hypothesis-generating candidates rather than generalizable findings.

In the present analysis, we tracked multiple sequelae within the same women across four omics modalities and repeated patient-reported outcomes, from a pre-treatment baseline through two years post-chemotherapy, each serving as her own control. Prior studies have typically evaluated a single exposure against a single outcome, such as endocrine assignment against weight^19,20^ or aromatase inhibitors against arthralgia,^21,39^ and those pairing molecular measurements with patient-reported outcomes have used limited panels at few timepoints.^17,49^ A systematic review of 34 such studies found inflammatory associations to be inconsistent in cross-sectional designs and more reproducible in longitudinal ones.^17,49^ Our sampling density lets us separate side effects by onset and distinguish the timelines of RAB37 and INHBB responses, and we offset the small sample size by validating both markers in the UK Biobank and restricting the microbiome analysis to three pre-specified butyrate producers. We propose that persistent adverse effects in breast cancer survivors, which differ in their timing of onset, are better understood as longitudinal trajectories than as a single post-chemotherapy state, and that these trajectories leave molecular traces months before the symptom itself is established. If such markers hold in larger cohorts, they would identify at-risk women during treatment, allowing supportive interventions limiting the sequelae that erode quality of life in survivorship.

## Methods

### VIBE study design and data collection

The VIBE study was a prospective longitudinal cohort conducted by the Institute for Systems Biology (ISB) and the Providence Swedish Cancer Institute (SCI). Eligible participants were women older than 18 years with newly diagnosed AJCC stage I, II, or III invasive breast cancer whose management plan included systemic chemotherapy, enrolled before any therapeutic intervention. Chemotherapy regimens included taxane and cyclophosphamide, doxorubicin and cyclophosphamide, and docetaxel-based regimens with or without trastuzumab **(Supplementary Table 2)**. Patients could move between adjuvant regimens and remain on study. Overall, 16 patients had evaluable longitudinal data and are analyzed here. Study visits were scheduled at screening and baseline, at the midpoint of chemotherapy, within one month of the last chemotherapy dose, and at 3, 6, 12, 18, and 24 months post-chemotherapy, with a visit window of ±2 weeks. Patient-reported outcomes were collected with FACT-B+4 (Functional Assessment of Cancer Therapy Questionnaire for Breast Cancer), FACT-Fatigue, PANAS (Positive and Negative Affect Schedule), and SF-12 (Short Form 12-Item Health Survey 2.0), together with the PRO-CTCAE (Patient-Reported Outcomes Version of the Common Terminology Criteria for Adverse Events) for treatment-related toxicity and dietary questionnaire, adapted from the Dietary Targets Monitor,^50^ together with internal questionnaires covering digestion, stress, and lifestyle. Blood was drawn eight to nine times per patient, after a 12-hour fast and before 11 am at every visit, so that circulating measurements are comparable within and across patients. Clinical blood chemistry (CBC with differential and comprehensive metabolic panel) and additional chemistry panels were run by LabCorp, metabolomics by Metabolon, and plasma proteomics with Olink immunoassays. Stools for gut microbiome profiling were collected at four to five time points using home kits returned at the following clinic visit, and waist and hip circumference were measured at each visit.

### Clinical and dietary data processing

For body measurements (weight, body temperature, systolic/diastolic blood pressure, respiratory rate, and heart rate), baseline values were subtracted from subsequent measurements for each patient. The resulting differences were divided by baseline measurements to calculate percent change for each parameter. BMI was calculated from weight and height for each patient and normalized in the same manner. For self-reported data, we selected 128 fields relevant to the frequency or severity of adverse effects out of 294 total fields. Ordinally categorized variables were converted into numeric values. For example, for severity: ‘*Never’*, ‘*Mild’*, ‘*Moderate’*, ‘*Severe’*, ‘*Very severe’* were assigned values of 1,2,3,4, and 5, respectively. For frequency: ‘*Never’*, ‘*Rarely’*, ‘*Occasionally’*, ‘*Frequently’*, ‘*Almost constantly’* were assigned values of 1,2,3,4, and 5. For variables such as “*moderate activities for at least 10 minutes*” and “*the frequency of stretching/flexibility activities*,” the following coding was applied: “*rarely or never*” = 1, “*less than once per week*” = 2, “*1-2 times per week*” = 3, “*at least 3 times a week*” = 4, and the highest two frequency levels (i.e., ≥5-7 times per week) = 5. Binary variables (Yes/No) were coded as 5 and 0, respectively. The stress score (1 = no stress, 10 = the most stress) was rescaled to a range of 0.5 to 5. Sleep duration was coded as follows: <4 hours = 1, 4-5 hours = 2, 6-7 hours = 3, 8-9 hours = 4, and >9 hours = 5. The normalized scored variables were then integrated into 41 side-effect events by averaging the variables belonging to the same side-effect category, accounting for the directionality of the effect. For example, for appetite, variables related to decreased appetite (“*Do you have a poor appetite and/or skip meals?*”, “*In the last 7 days, what was the severity of your decreased appetite at its worst?*”, “*In the last 7 days, how much did decreased appetite interfere with your usual or daily activities?*”) were multiplied by −1 before integration. Similarly, for sleep quality, variables indicating poor sleep (“*In the last 7 days, what was the severity of your insomnia-including difficulty falling asleep, staying asleep, or waking up early-at its worst?*”, “*In the last 7 days, how much did insomnia interfere with your usual or daily activities?*”) were multiplied by −1 before integration. Dietary intake was assessed by the Dietary Targets Monitor questionnaire.^50^ We first converted ordinally categorized intake variables into numerical values by creating scaled dummy variables ranging from 0 to 9, as follows. For food intake frequency: “*< Once per month*” = 1, “*1-3× per month*” = 2, “*Once per week*” = 3, “*2-4× per week*” = 4, “*5-6× per week*” = 5, “*Once per day*” = 6, “*2-3× per day*” = 7, “*4-5× per day*” = 8, and “*>6× per day*” = 9. For serving size per day: “*Zero/less than 1 per day*” = 0, “*1 serving*” = 3, “*2-3 servings*” = 6, and “*4-5 servings*” = 9. For probiotic supplements: “*Not at all*” = 0, “*Once per week or less*” = 3, “*Several times per week*” = 6, and “*Daily*” = 9. Dairy avoidance (*No*/*Yes*) was coded as 1 and 0, respectively. Next, the rescaled values belonging to the same food categories were averaged. For instance, frequency metrics for green, root, cruciferous, and salad vegetables were averaged alongside vegetable serving size to generate a composite "*vegetable*" variable. Similarly, variables representing the frequency of consuming chocolates, ice cream, cakes, scones, sweet pies, pastries, and other sweets were averaged to create a “*sweets*” variable. The variables for crackers, pretzels, chips, savory snacks, and other between-meal snacks were averaged to create a “*snacks*” variable. The frequency and serving size of juice consumption were combined into a “*juice*” variable. Variables for yogurt, fermented foods (e.g., sauerkraut, kombucha, kimchi), and probiotic supplements were averaged to create a “*probiotic food*” variable.

### Omics data processing

For clinical chemistries, analytes with more than 50% missing values across samples were excluded. Zero measurements were imputed using half the minimum observed non-zero value (i.e. 0.5*minimum) followed by log-transformation. Data were subsequently mean-centered across samples. For metabolomics (Metabolon), metabolites present in at least 80% of samples were retained. Similar to blood chemistry, raw measurements were log-transformed and mean-centered. For proteomics (Olink), Normalized Protein Expression (NPX) data were used. Proteins with more than 50% missing values were excluded.

### Gut microbiome data processing

Microbiome taxonomic signatures were estimated using a custom nextflow pipeline^51^. Initial quality control was performed using fastp^52^ (v0.24.0), where reads were trimmed to remove low-quality bases, with a minimum quality threshold set at 20, a minimum read length of 50 bp, and a maximum read length of 150 bp to ensure the retention of high-quality data. Taxonomic relative abundances were then estimated using MetaPhlAn (v.4.1.1)^53^ and reformatted into a tabular format for subsequent analysis.

### Statistical analysis of clinical and patient-reported outcomes

The time window from the start of chemotherapy to within 3 months after its end was defined as the chemotherapy period (Chemo), and the period from >3 to 24 months post-chemotherapy was defined as the post-chemotherapy period (PostChemo). The scores for each adverse effect, physical activity, and body measurement were baseline-centered by subtracting patient-specific baseline values. The statistical significance of score changes for each event or body measurement was assessed using a two-tailed, one-sample Wilcoxon signed-rank test. A median score value significantly different from zero indicated a significant change in the event during the specified period (Chemo or PostChemo).

### Statistical association analyses of clinical and longitudinal dietary predictors

To evaluate associations between baseline demographic and clinical features and long-term outcomes (weight gain and fatigue), we fitted GLMs using the brglmFit method.^54^ For the weight gain analysis, percentage weight changes at 24 months post-chemotherapy were used as continuous variables assuming a Gaussian error distribution. For the fatigue analysis, a binary outcome (fatigue vs. non-fatigue, defined by an outlying fatigue score increase at 24 months post-chemotherapy) was applied assuming a binomial error distribution. The following features were analyzed in separate GLM regression: age, weight, BMI (calculated as weight [kg]/height [m]^2^), menopausal status, breast cancer subtype (hormone receptor-positive [HR+], triple-negative [TNBC], or HER2-enriched [HER2+]), disease stage, and treatment types (neoadjuvant chemotherapy, endocrine therapy, endocrine therapy duration, mastectomy status, and total surgical interventions).

For longitudinal dietary data, association analyses across the 28 dietary parameters were performed using GEEs implemented via the *gee* package^30^ in R, with percent weight change or fatigue score as continuous response variables. Models specified a Gaussian error distribution and an independence correlation structure, with patient ID defined as the clustering subject variable. Two-tailed p-values were derived from the robust z-scores generated by the GEE models and FDR-corrected for multiple hypothesis testing.

### Fatigue omics signature analysis

Participants were classified into a fatigue group (n = 4, defined by severe fatigue at 24 months post-chemotherapy) and a non-fatigue group (n=8, representing the remaining participants); four individuals (Patients 109, 111, 117, and 118) were excluded from post-chemotherapy fatigue analyses due to missing 24-month data. To identify longitudinal multi-omics signatures (metabolomics, proteomics, and blood chemistry) during the period of post-chemotherapy (between 12-24 months after chemotherapy), binomial logistic regression was conducted using the *gee()* function in R, with an independent correlation structure.

### Evaluation of INHBB and RAB37 biomarkers across UK Biobank fatigue subgroups

We utilized two primary UK Biobank^31^ variables to evaluate INHBB and RAB37 expression levels: Chronic fatigue (Field 120114: persistent or recurrent tiredness lasting for at least 6 months) and Fatigue severity over the past week (Field 120040: mild, moderate, or severe). The reference (non-fatigue) group comprised individuals with no current mild or severe fatigue (Fields 28696, 28699) and no history of chronic fatigue, defined by a response of ‘*no problem*’ on Field 120040 and a negative report on Field 120114. Responses of ‘*Do not know*’ and ‘*Prefer not to answer*’ were excluded. Expression levels of INHBB and RAB37 markers were compared between each fatigue group and the reference group, using a two-sample t-test.

### Relative abundance and association analysis of *F. prausnitzii*

Relative abundances generated by MetaPhlAn (v.4.1.1)^53^ were calculated by normalizing individual species read counts against total read depth per sample. Logistic regression models were implemented using the *logistf*^55^ package in R to model fatigue status (0 = non-fatigue, 1 = fatigue) as a function of species relative abundance.

### Community-scale metabolic modelling and SCFA prediction

Steady state SCFA fluxes and microbial community growth rates were predicted using the community-scale metabolic modelling platform MICOM (v0.37.1) as described previously^56^. MCMMs^35^ were built using the species level taxonomic profiles and the MICOM *build()* function with a relative abundance threshold of 10^-3^, omitting taxa that made up less than 0.1% relative abundance. Metabolic models from the AGORA2 database^57^ (v2.01) summarized to the species level were used for taxa present in each MCMM. Steady-state growth rates and metabolic fluxes for all samples were then inferred using the MICOM *grow()* function with a tradeoff parameter of 0.8 and the “strategy=none” option. For all growth simulations a medium representing an average european diet with metabolites known to be absorbed in the small intestine depleted was utilized as a template and personalized for individuals based on their dietary questionnaire data. The questionnaire categories utilized for this purpose were ‘Beans or legumes (e.g. lentils, peas, pinto beans, etc.)’, ‘Cooked green vegetables (fresh or frozen)’, ‘Cooked root vegetables (fresh or frozen)’, ‘Cruciferous vegetables (e.g. broccoli, cabbage, cauliflower, etc.)’, ‘Fresh fruit’, ‘Nuts/seeds (e.g. almonds, walnuts, flax seeds, etc.)’, and ‘Raw vegetables or salad (including tomatoes)’. Consumption frequencies and category specific fiber averages per serving were used to estimate daily fiber intake for each sample. Estimated daily fiber intake was then used to scale the medium fluxes for a set of twenty-four dietary fibers present in the medium. The sample specific personalized diets were then utilized for growth simulations.

### Statistics and reproducibility

Due to the small sample size, statistical approaches were strictly selected to avoid parametric assumptions and prevent model overfitting. Differences between independent subgroups (e.g., fatigue vs. non-fatigue; short vs. long endocrine exposure) were evaluated using two-tailed Mann-Whitney U tests. Changes in self-reported side-effect scores and body measurements during and post-chemotherapy relative to baseline (Δ=x_chemo or post_−x_baseline_) were evaluated using two-tailed, one-sample Wilcoxon signed-rank tests against a null hypothesis of zero change (**Fig. 2**). Side-effect variables with a median change of zero were excluded from statistical testing to eliminate uninformative variables and prevent statistical artifacts driven by extreme sparsity. Non-parametric methods were chosen because they do not assume underlying Gaussian distributions and remain robust against leverage driven by extreme values in small sample sizes. The 10-month threshold separating short-term and long-term endocrine therapy exposure was derived empirically through data-driven exploration. Density distribution profiling across 12, 18, and 24 months post-chemotherapy revealed a bimodal separation with a distinct gap near 10 months. Because standard clinical thresholds (e.g., 5 years) exceeded our follow-up window, this pragmatic threshold was adopted to maximize biological group separation without imposing arbitrary cutoffs. To evaluate clinical variables and the gut microbiome, we fitted GLMs using bias-reduced estimation (*brglmFit* in the *brglm2* R package)^54^ and Firth’s penalized likelihood estimation (*logistf*)^55^, respectively. These methods modify the score functions to deliver finite, unbiased β coefficient estimates and reliable confidence intervals, avoiding non-convergence and inflated parameter estimates caused by small sample sizes and low event counts. To evaluate time-series dietary variables, we used GEEs with an independence correlation structure to prevent non-convergence and avoid model over-parameterization given our moderate sample size. For all statistical tests, exact sample sizes (n), exact P-values, and FDR-adjusted p-values (for multiple-hypothesis testing) are reported. Specific variable exclusions (e.g., metrics with a median change of zero) are explicitly noted. Finally, statistical analyses above were conducted using R (version 4.4.1).

### Use of large language models

The authors used a large language model (Claude, Anthropic) to assist with drafting and editing the manuscript text. All content was reviewed, verified, and edited by the authors, who take full responsibility for the work.

---

**Supplementary Figure 1.**
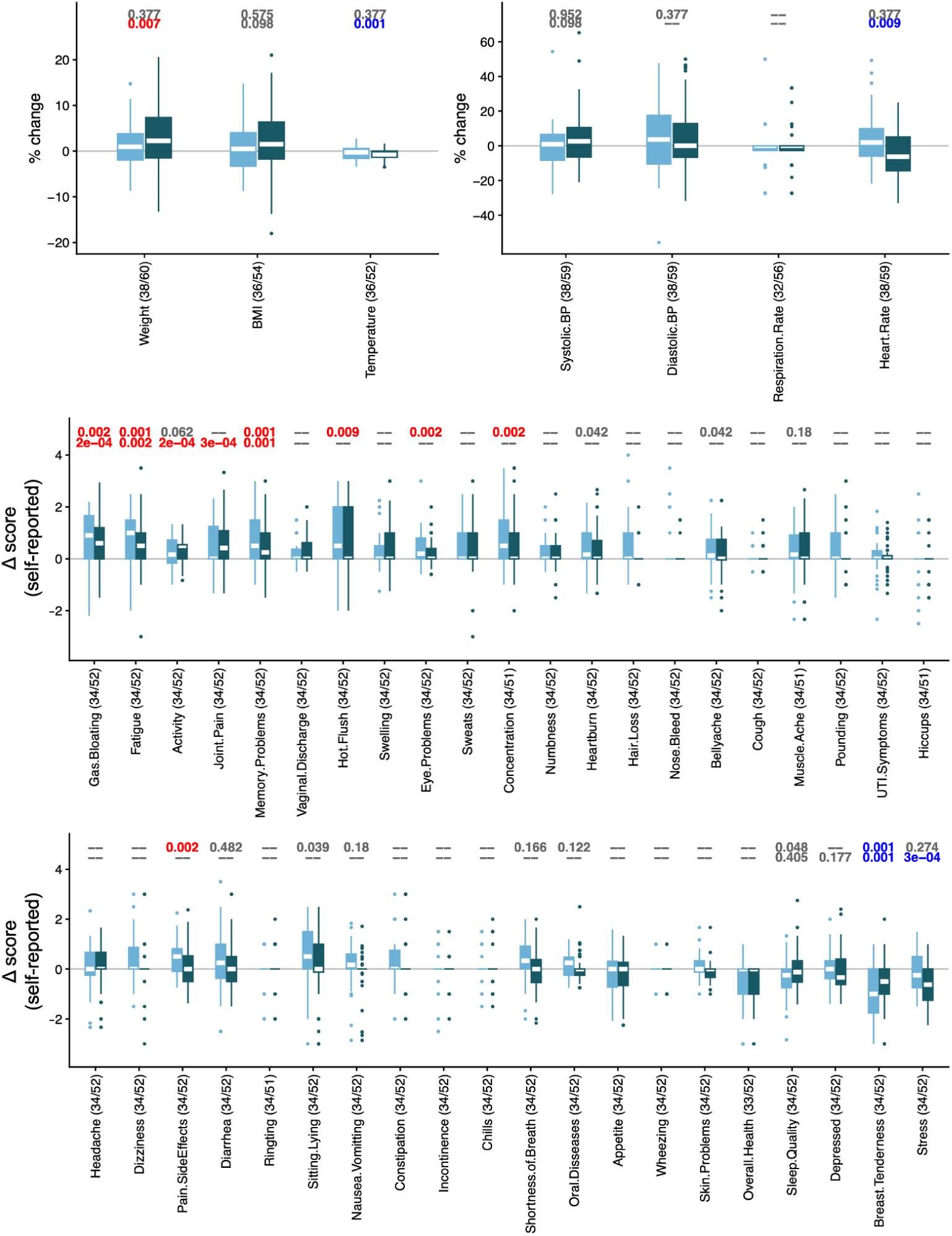
Top panels display percent changes in body measurements, while the last two panels display score changes across 41 self-reported side-effect groups relative to baseline during chemotherapy (light blue; treatment to 3 months post-last dose) and post-chemotherapy (dark blue; 3–24 months post-treatment). Side-effect scores were normalized (0–5 scale; Methods). Box plots show the median (center line), upper and lower quartiles (box limits), and whiskers extending to 1.5×IQR (interquartile range) from the hinges. Individual points beyond the whiskers denote outliers. Sample sizes for each test are annotated on the x-axis labels as "side effect (n[Chemo]/n[PostChemo])". Statistical significance was assessed using two-tailed, one-sample Wilcoxon signed-rank tests. Top annotations show FDR-adjusted p-values (q<0.01), where red denotes significant increases and blue denotes significant decreases. Variables with a median of 0 were excluded from statistical testing ("--").

**Supplementary Figure 2.**
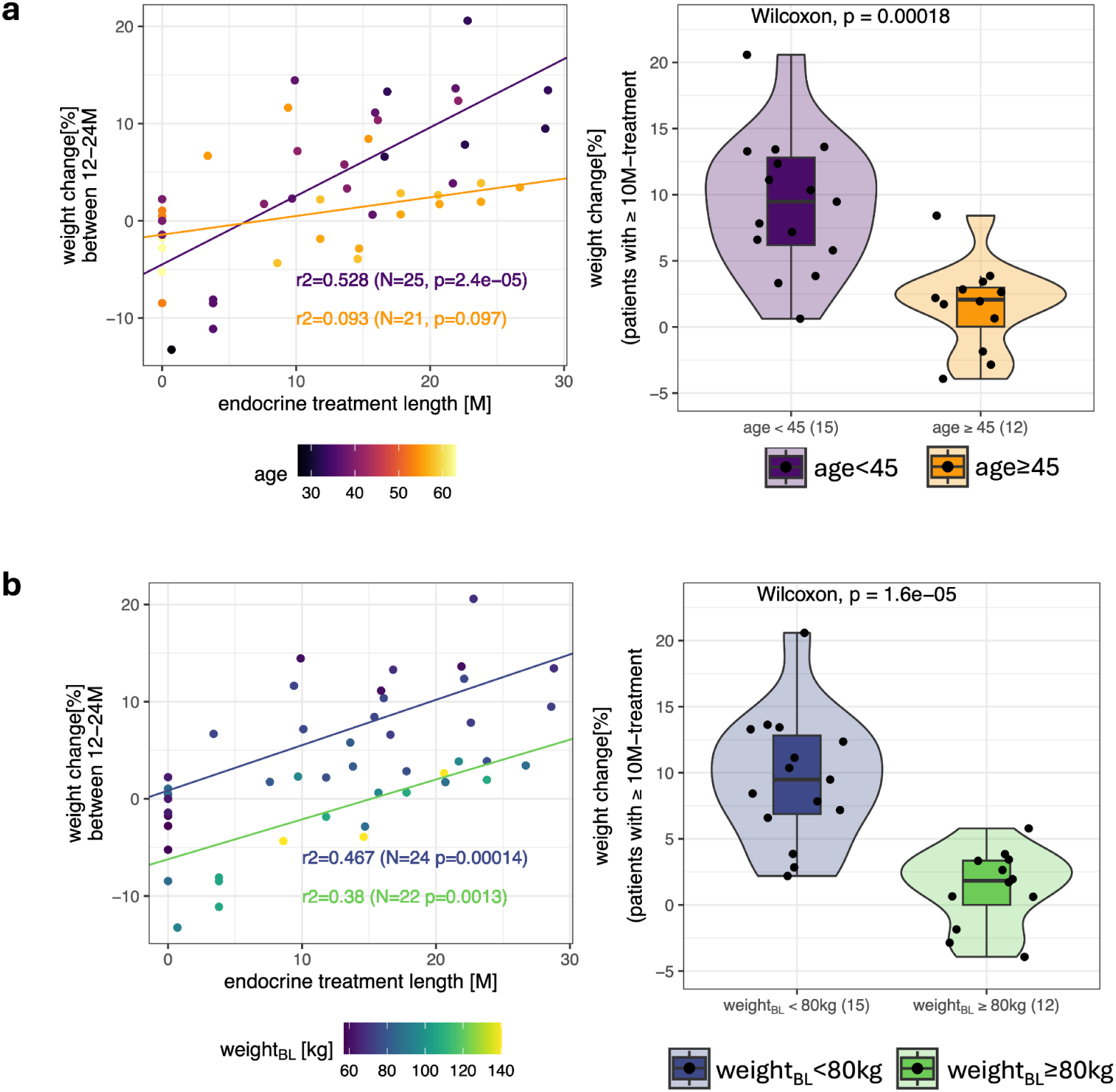
**(a, b)** Scatter plots displaying percent weight change against endocrine treatment duration (M, months), pooled across 12, 18, and 24 months post-chemotherapy. Data are colored by **(a)** age group and **(b)** baseline weight group, with linear regression fits, r^2^, sample sizes (N), and p-values indicated for each group. **(c, d)** Violin plots showing percent weight change between 12 and 24 months post-chemotherapy among patients who received ≥10 months of endocrine therapy, stratified by **(c)** age (< 45 years [purple] vs. ≥ 45 years [orange]) and **(d)** baseline weight (< 80kg [blue] vs. ≥ 80kg [green]). Group distributions were compared using two-tailed Mann-Whitney U tests. Violins display kernel density distributions with inset box plots and individual data points overlaid; box plots depict the median (center line), upper and lower quartiles (bounds), and whiskers extending to 1.5×IQR(interquartile range). Sample sizes (N) are indicated in parentheses on the x-axis labels.

**Supplementary Figure 3.**
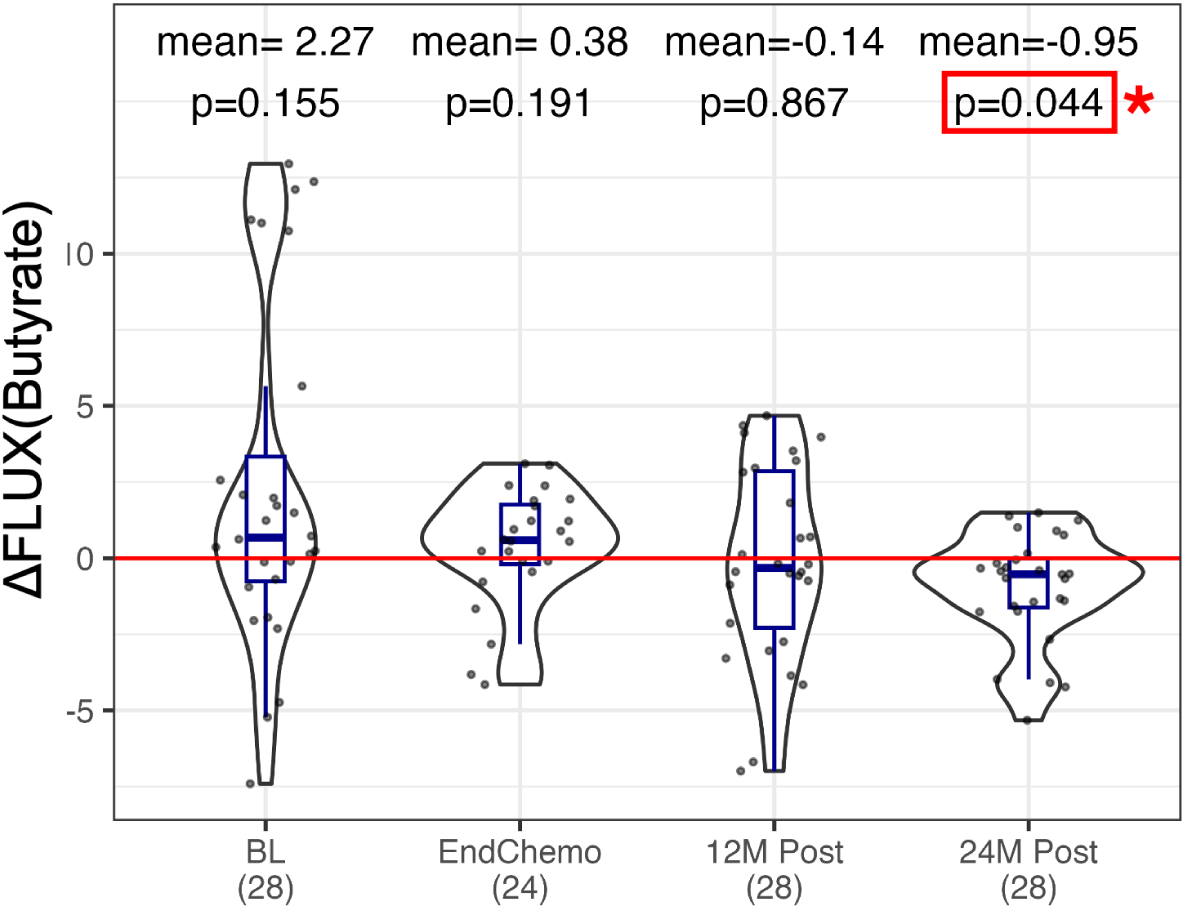
Differences in estimated butyrate fluxes (Δflux = Butyrate_fatigue_ − Butyrate_non-fatigue_) between the fatigue and non-fatigue groups across four timepoints: Baseline (BL), end of chemotherapy (EndChemo), and 12 months (12M Post) and 24 months (24M Post) post-chemotherapy. Pairwise flux differences were generated using permutations within each timepoint. Box plots show the median (center line), upper and lower quartiles (box limits), and whiskers extending to 1.5×IQR(interquartile range) from the hinges. Individual data points are overlaid. FDR-adjusted p-values from two-tailed, one-sample Wilcoxon signed-rank tests as well as mean Δflux are annotated above each box plot, and sample sizes (N) are reported on the x-axis labels as "Time (N)". Statistically significant values (p<0.05) are highlighted with a red outline.

**Supplementary Table 1.**

| **UK Biobank** |  | **INHBB** |  | **RAB37** |  |
| --- | --- | --- | --- | --- | --- |
| length of experiencing mild fatigue | n | t-value | p-value | t-value | p-value |
| < 2 weeks (ref.) | 654 | 0.00 | 1.0000 | 0.00 | 1.0000 |
| 2-4 weeks | 641 | 0.69 | 0.4930 | 1.61 | 0.1085 |
| 4-12 weeks | 1168 | 1.37 | 0.1723 | -0.03 | 0.9768 |
| >12 weeks | 5026 | 2.62 | 0.0090 | -0.48 | 0.6284 |
| length of experiencing severe fatigue | n | t-value | p-value | t-value | p-value |
| < 2 weeks (ref.) | 18 | 0.00 | 1.0000 | 0.00 | 1.0000 |
| 2-4 weeks | 16 | 0.99 | 0.3315 | -0.68 | 0.5040 |
| 4-12 weeks | 21 | 1.17 | 0.2540 | -1.61 | 0.1261 |
| >12 weeks | 193 | 1.71 | 0.1064 | -0.56 | 0.5799 |

## Author contributions

E.D.E., T.B., N.D.P., and A.T.M. conceived and designed the study cohort, with input from L.H. during protocol development. E.D.E. served as principal investigator and oversaw the clinical conduct of the study. K.M. provided follow-up clinical care. S.M.M. and D.B. coordinated patient care, study visits, and sample collection. N.B. provided research administration and regulatory oversight at the Providence Swedish Cancer Institute. M.E.B. managed study operations and data curation at ISB. A.V.C. and N.Q.-B. performed metagenomic processing and community-scale metabolic modelling. H.N. and L.P. performed the statistical and multi-omic analyses. S.M.G. supervised the microbiome analyses. C.W.D. contributed to clinical interpretation. A.T.M. and N.R. supervised the analysis and interpretation. H.N. and N.R. wrote the manuscript. All authors reviewed, edited, and approved the final manuscript.

## Acknowledgements

This study was funded by the Swedish Foundation, and a generous gift from K. C. Ellison (to H.N.). We are grateful to Mariko Tameishi, Evonne Lackey and Jef St. DeLore (Swedish Cancer Institute) for program coordination and regulatory oversight; Jeff Boore, Ramkumar Hariharan, Max Robinson (ISB) for contributions during early stages of project planning. And we are especially grateful to the patients who selflessly gave their time and energy to this study.

## Data and Code availability

Qualified researchers can access the de-identified VIBE dataset for research purposes through a Data Use Agreement. Inquiries about data access should be sent to and will be responded to within seven business days. Access to UKBB data is subject to approval and can be requested through their application process. The UKBB approved the use of data for this study under application number 48049. Code used in this study is freely available on GitHub (https://github.com/PriceLab/VIBE).

